# Comparative evaluation of imputation and batch-effect correction for proteomics/peptidomics differential-expression analysis

**DOI:** 10.1101/2025.08.14.25333694

**Authors:** Charis Gonidaki, Agnieszka Latosinska, Antonia Vlahou, Rafael Stroggilos, Harald Mischak

**Author notes:** **Corresponding authors:** Harald Mischak, Mosaiques Diagnostics GmbH, Rotenburger Str. 20, D-30659 Hannover, Germany; Charis Gonidaki, Department of Biotechnology, Biomedical Research Foundation of the Academy of Athens (BRFAA), 4 Soranou Efessiou Str., Athens 11527, Greece.

## Abstract

Mass spectrometry (MS)-based proteomics offers powerful opportunities for biomarker discovery; nevertheless, it is associated with technical challenges, some of them being missing values and batch effects. Both can obscure biological signal and bias results. Although imputation and batch-correction methods are well established in transcriptomics, their impact, particularly on large-scale, real-world clinical proteomics datasets, remains unclear. In this study, we systematically compared the impact of two popular imputation methods (½ LOD replacement and KNN) in combination with three batch-effect correction approaches (ComBat, ComBat with disease covariate, and MNN) on differential expression analysis in a CE-MS urine peptidomics dataset of 1,050 samples across 13 batches collected for early detection of chronic kidney disease (CKD), separated into discovery (n = 525) and validation (n = 525) sets. Our results show that the choice of imputation method (between ½ LOD and KNN) had minimal impact on the final list of differentially expressed peptides (DEPs). In contrast, batch-effect correction had a much stronger influence on the results. ComBat without covariate adjustment removed most DEPs, suggesting loss of true biological signal. Along these lines, incorporating disease status into the model preserved most of this information. MNN yielded a moderate to low number of validated DEPs overall, especially when paired with KNN imputation. These findings show that imputation and batch correction are not entirely independent processes and that they can influence downstream results. Overall, preprocessing choices should be chosen based on the characteristics of each dataset and especially considering batch severity and biological covariates.

**Statement of significance of the study:** Finding reliable biomarkers in clinical proteomics first requires addressing the technical noise that can hide true biological signals. In this work, we investigate how different imputation and batch correction methods influence the list of peptides that emerge as differentially expressed. Instead of relying on simulations or small datasets, we examine a large, real-world urine-peptidomics cohort of more than 1,000 samples screened for early-stage chronic kidney disease. The results show that no preprocessing pipeline is universally optimal and that the best choice depends on the characteristics of the dataset. This study offers practical guidance for improving reproducibility in urine-based peptide studies and supports more confident identification of disease-associated molecular signatures.

## 1. Introduction

Mass spectrometry (MS) has become one of the most powerful tools in biomedical research for detecting and quantifying proteins and peptides in biological samples. Over the past decade, major advancements in MS instrumentation, including higher resolution, enhanced sensitivity, and faster acquisition rates, have greatly improved its performance^[1]^. These improvements have allowed researchers to detect low-abundance proteins and peptides, resolve closely related molecular species, and analyze thousands of features across large sample cohorts. As a result, MS-based platforms now play a central role in biomarker discovery, disease classification and personalized medicine^[2]^.

Despite these advances, proteomics, in fact omics approaches in general, face several technical challenges. Among the most prominent are missing values and batch effects. Missing values leave gaps in the data that can skew statistical analyses. Their causes fall into three main categories^[3]^,^[4]^. If a feature’s intensity is lost purely by chance (e.g., a transient instrument glitch), it is considered missing completely at random (MCAR). When detection depends on other measured factors (such as generally low signal across runs) but not on the feature’s own true abundance, data are missing at random (MAR). Finally, features with true concentrations below the detection limit produce missing not at random (MNAR) values. Although these mechanisms, in practice, often overlap, distinguishing among them guides the choice of imputation strategy^[5]^.

In parallel, batch effects introduce non-biological variation when samples are collected at different centers (introducing center bias) or processed on different days, with different instruments, or in different laboratories. These effects can similarly obscure genuine biological signals^[6]^.

Over the years, many imputation and batch-correction methods have been developed in order to address these distinct sources of variation. Two of the most widely used imputation techniques are half-LOD (½ LOD) replacement, which is well suited for MNAR scenarios, and K-nearest neighbors (KNN), which can handle MAR patterns by borrowing information from similar features^[7]^,^[8]^. Imputation is also commonly applied as a preparatory step, since many downstream methods such as batch correction assume complete data and cannot directly handle missing values. Likewise, batch-correction approaches like the empirical-Bayes ComBat method and the mutual nearest neighbors (MNN) algorithm each make different assumptions about how non-biological variation should be modeled and removed^[9]^,^[10]^.

Numerous benchmarking studies independently evaluate imputation tools^[8]^,^[7]^ or batch-correction algorithms^[11]^,^[12]^, yet only a handful have attempted to consider how the two steps interact, or how those interactions affect downstream differential expression analyses^[13]^. Most studies apply a single preprocessing pipeline, leaving it unclear which combinations best remove technical noise while preserving biological signals^[14]^.

To address this gap, we utilized peptidomics data from samples collected from 13 different research hubs (batches) and performed a comprehensive, pairwise evaluation of two imputation approaches (½ LOD replacement and KNN) and three batch-effect correction methods (ComBat, ComBat with the disease status as a covariate and MNN). These summed for a total of 8 different processing pipelines which were assessed alongside an unmanipulated baseline dataset (no imputation and no batch-effect correction).

As a test case, we used urine peptide data collected via capillary electrophoresis mass spectrometry (CE-MS), in the context of chronic kidney disease (CKD). The readout chosen was peptides differentially expressed (DEPs) between cases and controls. To evaluate the performance of each preprocessing strategy, we assessed both the consistency of DEPs between discovery and validation sets, and the reproducibility of DEPs under different preprocessing scenarios.

## 2. Methods

### 2.1. Dataset Description

An anonymized urine-peptidomics dataset was obtained from the Mosaiques Diagnostics database. It contained data from 1,224 samples (612 healthy controls, 612 Chronic Kidney Disease (CKD) patients) drawn from 36 independent studies (batches). Each sample included measurements for 24,346 unique Peptide IDs, acquired using capillary electrophoresis mass spectrometry (CE-MS).

For each of the 1,224 samples, clinical data were also available. In particular, the disease status (healthy control vs. CKD), sex (male/female), age (years), estimated glomerular filtration rate (GFR, mL/min/1.73 m²), diastolic blood pressure (dBP, mmHg) and systolic blood pressure (sBP, mmHg) were recorded.

As reliable imputation and batch correction require sufficient sample sizes, any study (batch) with fewer than 20 samples was excluded. This reduced the dataset to 13 batches, corresponding to 565 controls and 500 CKD cases. The retained batches ranged in size from 21 to 332 samples. An overview of the sample composition and data quality across the 13 batches is provided in ***Supplementary Table S1*.**

After matching individuals based on age, sex, and systolic/diastolic blood pressure, the final number of samples included in the analysis was 565 for the control group and 485 for the CKD patient group (1,050 samples in total). For downstream analysis, the samples were randomly split into discovery and validation subsets of equal size (n = 525 each), ensuring a balanced distribution of clinical and pathological variables (***Supplementary Table S2)***.

Peptides were further filtered based on the availability of sequence information, (reducing the total to 18,035 Peptide IDs) and a frequency threshold (peptides present in at least 50% of samples) which resulted in a final number of 3,262 Peptide IDs included in the analysis (***Supplementary Table S3)***. A concise overview of the final, filtered cohort is given in Error! Reference source not found..

### 2.2. Different combinations of methods used

Two imputation methods were tested individually and in combination with three batch-effect correction methods, resulting in eight distinct preprocessing pipelines.

#### Imputation methods

- **½ Limit of Detection (1/2 LOD),** which sets all missing values to half the minimum observed intensity in the dataset. This approach assumes that missingness arises because true peptide abundances fall below the instrument’s detection threshold^[15]^.
- **K-Nearest Neighbors (KNN),** this method replaces a missing value by averaging the values of the *k* most similar features, where “similarity” is defined via a chosen distance metric^[16]^.

In this study, we applied *VIM::kNN()* function to our peptide-intensity matrix, using k = 5 and using package’s default Gower type distance (which scales numeric variables to [0,1] before computing Euclidean distance)^[17]^.

#### Batch correction methods

The imputed datasets were log -transformed and three batch-correction variations were applied:

- **ComBat,** which applies either parametric or non-parametric empirical Bayes frameworks to adjust data for batch effects^[18]^. For this analysis, we applied the parametric empirical Bayes framework (package default) using the *`sva::ComBat()`* function on our log - transformed imputed peptide matrix. No biological covariates were included, so all between-batch variations were subject to removal.
- **ComBat using the CKD covariate,** which extends the above method by supplying a design matrix with the disease status covariate^[10]^.In this way, ComBat preserves differences associated with CKD status while still removing unwanted batch effects, ensuring that true case–control signal is not inadvertently washed out.
- **Mutual Nearest Neighbors (MNN),** this method identifies mutual nearest neighbors between datasets to align their shared structure^[19]^. These neighbors are pairs of data points (samples) from different batches that are closest to each other in the high-dimensional space, representing similar biological states. For this, we applied the function *`batchelor::batchCorrect()`* to our log -transformed imputed peptide matrix with *`PARAM = ClassicMnnParam(k = 15)`*, and *`cos.norm.out = FALSE`*.

### 2.3. Principal Components Analysis (PCA)

Principal Component Analysis (PCA) was performed on the log -transformed peptide-intensity matrices before and after batch correction. PCA plots with the first two principal components (PC1 and PC2) were generated using R to visually inspect the presence and strength of batch effects and to evaluate the extent to which correction methods reduced the unwanted variation.

### 2.4. Statistical Analysis

Differential expression analysis was performed as follows: To assess the stability of results, we applied the analysis separately to the previously defined discovery and validation sets (n = 525 each), which had balanced clinical and pathological characteristics. Within each subset, we performed non-parametric Mann–Whitney U-tests to compare CKD and control samples, followed by Benjamini–Hochberg adjustment to control the false discovery rate. Only peptides that were significant (adjusted p-value < 0.05) in both sets and showed the same direction of change were retained.

To further investigate the overlaps of the differentially expressed peptides and obtain an overview of the results, we calculated the Jaccard similarity index between all the different pairs of pipelines. The Jaccard similarity index measures the proportion of shared peptides between two sets relative to their total combined peptides. In mathematical terms the Jaccard index between two sets *A* and *B* is defined as^[20]^,

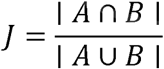

In our case, |A ∩ B| was the number of peptides that were found significant by both pipelines and |A ⋃ B| was the union of the significant peptides of these two pipelines. A higher *J* indicates greater overlap in significant peptides between the two methods.

## 3. Results

The goal of the study was to evaluate how different imputation methods and their pairwise combinations with batch-effect correction methods affect differential-expression analyses in urine peptidomics. The urine peptide dataset after filtering consisted of 3,262 Peptide IDs and 1,050 samples across 13 batches-studies.

### 3.1. Sample Distribution Across Pre-processing Pipelines

An unmanipulated “do-nothing” dataset, in which all missing values were set to zero and no batch correction was applied, served as the comparator. Seven core preprocessing pipelines: baseline (unmanipulated), ½ LOD-only, KNN-only, and each imputation paired with ComBat or MNN were employed.

Principal Component Analysis (PCA) was performed on the log -transformed peptide-intensity matrices before and after batch correction. **Figure 1** displays PC1 versus PC2 for each pipeline. As expected, the PCA results for the unmanipulated and ½ LOD imputed data were essentially identical (***Supplementary Figure S1***, **Figure 1A**). The KNN imputed data (**Figure 1B**) likewise show similar distribution of samples to the ½ LOD imputed data (**Figure 1A**). Therefore, simple imputation alone has almost no effect on the global variance structure, where samples from the same batch tend to cluster closer to each other. Notably, the degree of batch clustering in these imputation only panels appears relatively mild, suggesting that without explicit correction the inter study effect even though present is not overwhelming.

**Figure 1.**
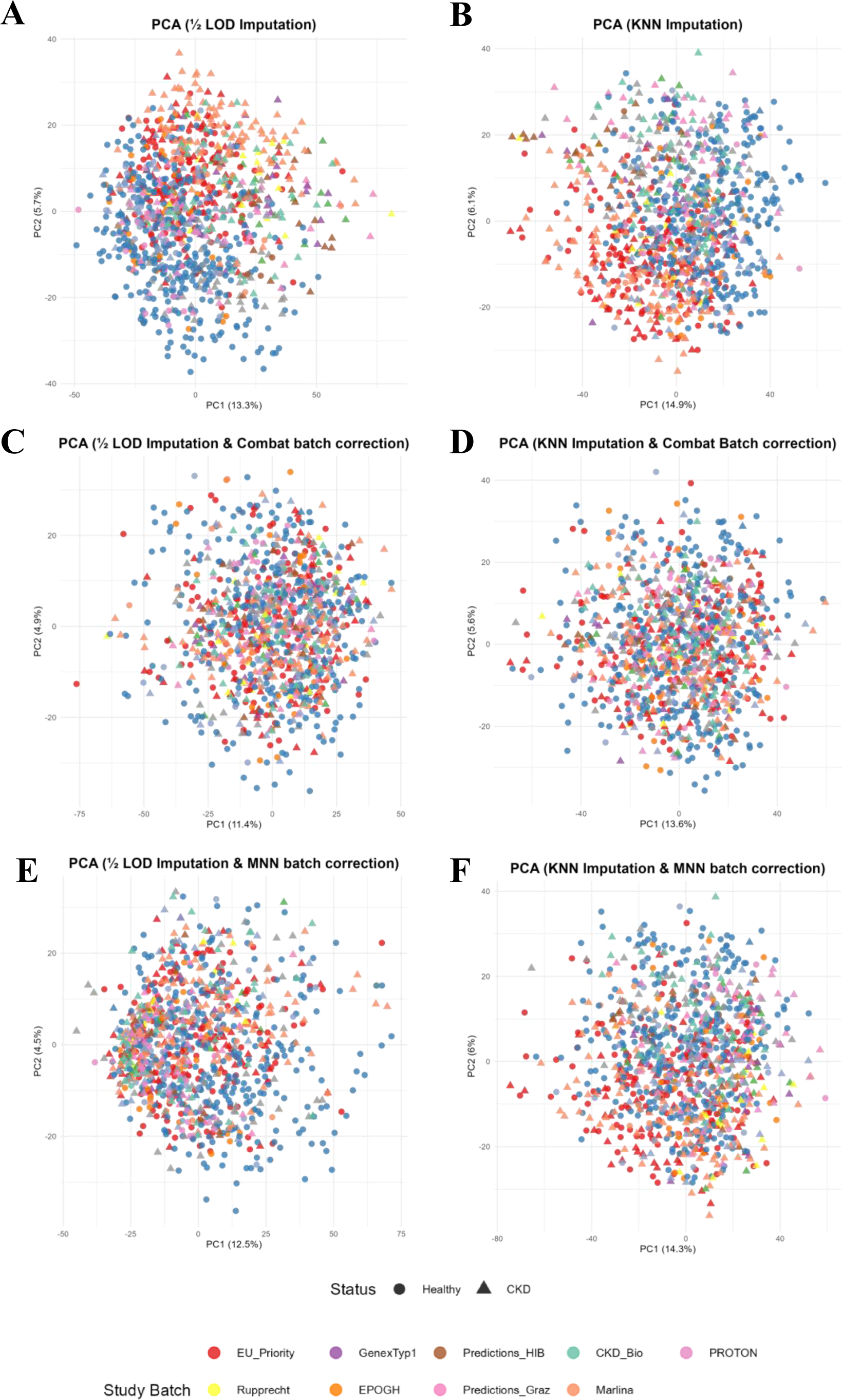
Principal Components Analysis (PCA) plots (PC1 vs. PC2) for six pipelines: **A)** ½ LOD, **B)** KNN, **C)** ½ LOD + ComBat, **D)** KNN + ComBat, **E)** ½ LOD + MNN, **F)** KNN + MNN. Each point represents a single sample, colored by its study batch and shaped by clinical status (● healthy, ▴ CKD).

In the batch corrected panels (**Figure 1C - F**), samples appear more evenly mixed, with no single study dominating any region. Yet, the shapes denoting clinical status also appear diffusely scattered, indicating that true disease related variance may have been reduced as well.

### 3.2. Impact of imputation and batch effect removal on significant peptides

Differential expression between CKD and healthy samples was assessed using Mann–Whitney U-tests with Benjamini–Hochberg correction. Only peptides that were significant in both the discovery and validation sets (with the same direction of change) were retained *(See* ***Supplementary Table S4*** *for the detailed results of the statistical analysis)*. The table in **Figure 2 A** summarizes the results of this analysis specifically depicting:

a. the total number of peptides that reached significance in the discovery set
b. the number of significant peptides replicated in the validation set with the same direction of regulation
c. the rate of discovery-set hits that were successfully validated.

**Figure 2.**
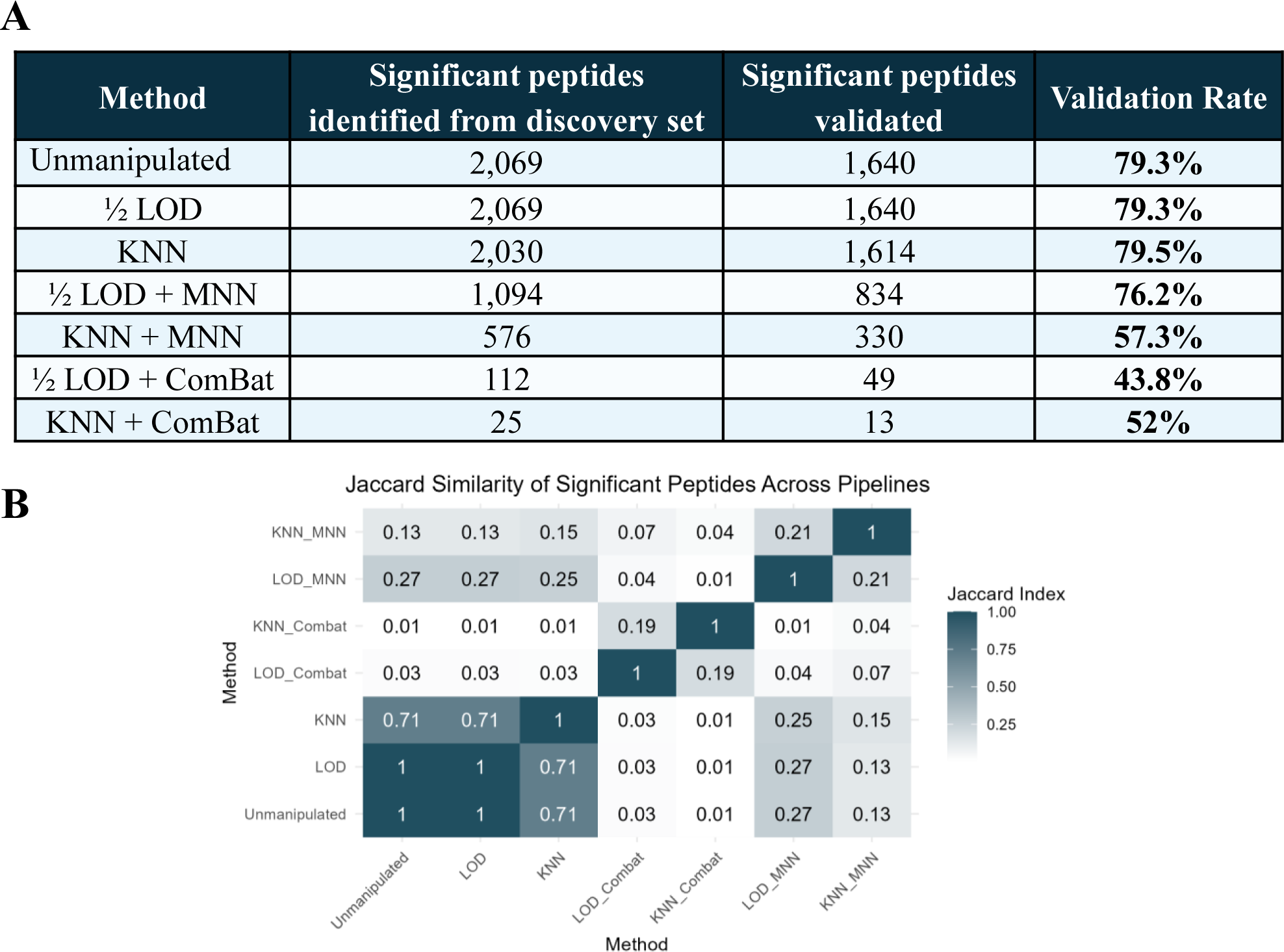
**A)** Table that, for each pipeline, reports the count of peptides found significant in the discovery set, the number of them that were validated in the validation set (with the same regulation trend) and the percentage of discovery-set hits validated. **B)** Heatmap showing the Jaccard index for the validated significant peptides, computed for every pair of the 7 core pipelines (unmanipulated, ½ LOD, KNN, and their combinations with ComBat and MNN). Each cell’s value and color intensity represent the fraction of shared significant peptides (darker = greater overlap).

As expected for a rank-based differential-expression test, replacing missing values with a constant (½ LOD) does not alter the relative ordering of peptide intensities. Consequently, the unmanipulated baseline and the ½ LOD-only imputed pipeline produced identical lists of significant peptides (1,640 validated peptides with validation rate 79.3%). KNN imputation resulted in a similar number of validated hits and validation rate (*χ*^2^= 0.004, p = 0.948). Adding batch correction uniformly and significantly reduced the number of significant peptides (from a mean of 1,630 before to fewer than 850 after batch-effect removal). The pipeline combining ½ LOD imputation followed by MNN batch correction (hereafter ½ LOD + MNN) retained a moderate set of validated peptides (n ≈ 830) while maintaining a high validation percentage (∼76%); these numbers declined further with KNN + MNN (n=330, < 60%). The drop was even more pronounced with ComBat, regardless of the imputation method applied (n ≤ 50, ≤ 52%).

Overlaps in findings of the different pipelines were further investigated using the pairwise Jaccard similarity index. The resulting values of the validated DEPs are displayed in Error! Reference source not found. **B**, where each cell’s color intensity reflects the degree of overlap between two pipelines (darker = greater overlap).

As evident from the heatmap, the unmanipulated baseline, ½ LOD and KNN pipelines share a high proportion of differentially expressed peptides (J > 0.7), reflecting similar results when no batch correction is applied. The two MNN pipelines (½ LOD + MNN and KNN + MNN) show low concordance with the other pipelines (J ≈ 0.15–0.3). This low index value is to a good extent expected, since the total number of significant peptides highlighted by pipelines using MNN is also lower. On the other hand, ComBat shows no considerable overlap with any pipeline (J < 0.1), highlighting its large impact on the dataspace, leading to the hypothesis that this approach may be removing not only batch-related variation but also biological signal.

To further investigate the hypothesis that ComBat removes biological variability, the impact of the algorithm was reapplied with the disease status (healthy control or CKD) included as a covariate (*see* ***Supplementary Figure S2*** *for the PCA plots*). Differentially expressed peptides (DEPs) were defined as previously: The Mann–Whitney U test with Benjamini–Hochberg correction was applied and only peptides found significant in both discovery and validation sets with the same regulation were retained

As shown in the table in **Figure 3 AFigure 3 A)** Table that, for each pipeline, reports the count of peptides found significant in the discovery set, the number of them that were validated through the validation set and the percentage of validated discovery-set hits. **B)** Heatmap that displays the Jaccard index for the validated DEPs calculated for every pair of nine pipelines (unmanipulated, ½ LOD, KNN, and their combinations with ComBat, ComBat(+CKD), and MNN). Each cell’s value and color intensity indicate the proportion of shared significant peptides (darker = greater overlap)., in both ½ LOD + ComBat(+CKD) and KNN + ComBat(+CKD) pipelines, a substantial number of DEPs were identified and successfully validated. Specifically, over 800 DEPs were detected in each pipeline, with validation rates around 75%. These results already show a significant increase in both the number and the percentage of validated differentially expressed peptides in comparison to the approach without the disease status as covariate.

**Figure 3.**
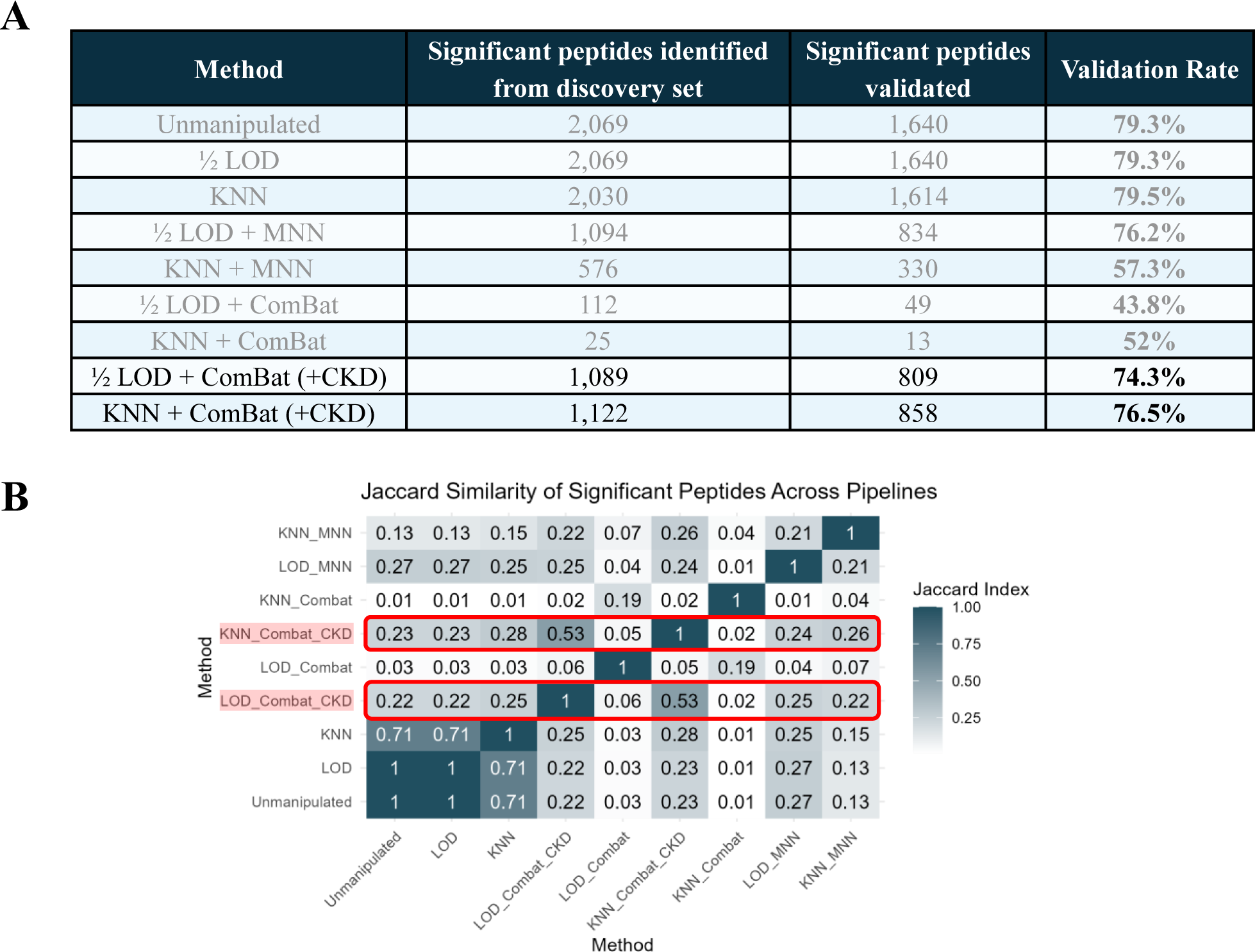
**A)** Table that, for each pipeline, reports the count of peptides found significant in the discovery set, the number of them that were validated through the validation set and the percentage of validated discovery-set hits. **B)** Heatmap that displays the Jaccard index for the validated DEPs calculated for every pair of nine pipelines (unmanipulated, ½ LOD, KNN, and their combinations with ComBat, ComBat(+CKD), and MNN). Each cell’s value and color intensity indicate the proportion of shared significant peptides (darker = greater overlap).

Additionally, to assess how including the CKD status as a covariant affects the statistical output, the Jaccard similarity index was calculated for every pair as previously (Error! Reference source not found. **B**). Both ComBat(+CKD) pipelines exhibit moderate overlap (J ≈ 0.2–0.55) with the rest methods, far exceeding the near-zero concordance seen using standard (unadjusted) ComBat. This heatmap further supports the hypothesis that ComBat without covariate adjustment removes biological variability.

## 4. Discussion

Even though analytical technologies have rapidly evolved, achieving remarkable progress in protein resolution, certain technical challenges remain difficult to overcome. Two persistent sources of variation that can bias data interpretation in -omics experiments remain the missing values and batch effects. This has led to the development of a range of imputation and batch-effect correction methods; however, the impact of these methods appears not too well known.

In this study, we evaluated how different combinations of widely applied imputation and batch-effect correction methods influence differential expression results in a large urine peptidomics dataset. We found that simple imputation methods (½ LOD or KNN) had minimal effect on the list of validated differentially expressed peptides (DEPs), closely matching the unmanipulated baseline. In contrast, batch-effect correction had a highly significant influence: ComBat without covariates drastically reduced the number of validated DEPs and had minimal overlap with other pipelines, while including disease status (CKD) in the model restored this signal to some extent. MNN provided an intermediate outcome, yet significantly reducing the ability to identify disease-associated biomarkers. When MNN followed KNN imputation the validated set size further decreased, indicating that the choice of imputation directly affects the performance of subsequent batch correction.

These findings show that the effects of imputation and batch correction are not independent and can interact in ways that affect downstream analyses. In our dataset, where batch-related variation was relatively mild, strong correction methods like unguided ComBat removed meaningful biological signals, which can be avoided by including biological covariates in the model or applying milder corrections like MNN. Therefore, it is important to let the structure of the dataset guide preprocessing decisions, especially considering how strong the batch effect is and whether relevant biological information is available. In well-balanced, multi-batch datasets like ours, minimal preprocessing may provide a good balance between keeping the biological signal and reducing technical variation. This limited batch-related variation may be explained by the normalization routinely applied to CE-MS datasets, which uses 29 collagen peptides as internal standards to harmonize peptide intensities across runs^[21]^,^[22]^.

The preprocessing methods examined in this study represent some of the most commonly applied approaches for addressing missing values and batch effects in omics research. Although their performance has been well studied, their impact on large-scale proteomics and peptidomics datasets is still not too well understood.

½ LOD imputation is commonly used in proteomics, especially when values are assumed to be missing not at random (MNAR) due to low abundance below the detection threshold. It is simple, fast, and preserves data structure, but it artificially reduces variance and can lead to biased fold-changes^[5]^,^[7]^. KNN imputation, by contrast, assumes missing at random (MAR) values and estimates them based on the similarity of peptides across samples. KNN is more flexible but it can over-smooth biologically relevant variation and it is sensitive to parameter settings^[7]^, ^[8]^. In our study, both methods produced similar results, indicating that the missing values were mostly MNAR, consistent with earlier findings in label-free proteomics^[23]^.

ComBat, a popular batch-effect correction method originally developed for microarray data, has also been widely adopted in proteomics due to its apparent robustness in small-sample settings. It applies empirical Bayes shrinkage to remove non-biological variation, but when the biological groups overlap with the batch structure, it risks removing true biological signal^[24]^, ^[10]^. MNN corrects batch effects by aligning shared structure across datasets using local neighborhoods, preserving more complex biological variation^[9]^. However, if batches are very small or uneven in size, MNN may compress the dataset’s dimensionality, which can weaken its statistical power^[9]^. This limitation may also be relevant in our dataset, given the variation in batch sizes.

Although these methods are widely used and supported by most analysis platforms, we acknowledge that more advanced imputation and batch correction approaches have been developed, especially in recent years. For example, several deep learning methods have shown promising results in both transcriptomics and proteomics^[25]^,^[26]^. In the context of imputation, other recent developments include statistical frameworks that either avoid imputation altogether or account for the variability it introduces into downstream analyses^[27]^,^[5]^. In this study, we chose to keep it simple and to focus on well-known and user-friendly methods that are commonly used in practice and easy to implement.

Overall, our findings reinforce the need to carefully evaluated any preprocessing step against the raw data, as such manipulations carry the risk of introducing artefacts and potentially distorting biological signals, ultimately preventing, rather than supporting the identification of significant features. Adaptation to the dataset’s specific characteristics, with attention to both technical variation and biological relevance, is essential.

## Concluding Remarks

In this urine-peptidomics dataset, the imputation of missing values (whether with ½ LOD or KNN) had little influence on the differential-expression analysis results. Batch-effect correction, on the other hand, reduced the number of peptides identified as significantly associated with the disease (CKE) without providing any evidence for improvement. Unguided ComBat removed most of the disease signal, and even less impactful approaches like MNN or CKD-guided ComBat led to significantly smaller DEP sets than the baseline. Overall, none of the tested preprocessing steps resulted in increased performance compared to the unmanipulated data. This may be linked to the normalization routinely applied to CE-MS datasets, based on 29 collagen peptides used as internal standards, which seems to minimize batch-related peptide variation.

These findings also confirm that imputation and batch correction do not act independently, and that no single preprocessing pipeline is universally optimal. The choice of whether and how to apply batch correction should depend on the strength of batch effect and the inclusion of relevant biological variants in the model. In well-balanced multi-batch datasets like ours, a minimal approach that focuses only on addressing missing values (and be it replacing missing values with 0) may be sufficient to retain meaningful biological signals and avoid unnecessary loss of information. Overall, this highlights the importance of tailoring preprocessing strategies to the specific characteristics of each dataset, rather than relying on a fixed pipeline for every dataset.

## Data Availability

All data needed to reproduce the results of this study are provided in the accompanying Supporting Information files.

## Abbreviations

CKD: Chronic Kidney Disease
dBP: Diastolic Blood Pressure
DE: Differential Expression
DEPs: Differentially Expressed Peptides
GFR: Glomerular Filtration Rate
J: Jaccard Similarity Index
KNN: k-Nearest Neighbors
MAR: Missing at Random
MCAR: Missing Completely at Random
MNAR: Missing Not at Random
MNN: Mutual Nearest Neighbors
sBP: Systolic Blood Pressure

## Supporting Information

**Supplementary Table S1** Overview of sample composition and data quality across the 13 studies.

**Supplementary Table S2** Sample-level metadata original study label, discovery/validation phase, and CKD status for all 1,050 urine samples analyzed.

**Supplementary Table S3** Peptide abundance matrix for all 1,050 urine samples analyzed.

**Supplementary Table S4** Full Detailed statistical results for all peptides.

**Supplementary Figure S1** Principal Components Analysis (PCA) plots (PC1 vs. PC2) for Unmanipulated and ½ LOD datasets.

**Supplementary Figure S1** Principal Components Analysis (PCA) plots (PC1 vs. PC2) for ComBat(+CKD) pipelines

## Acknowledgements

This article/publication is based upon work conducted within a Short-Term Scientific Action from COST Action CA21165 (PerMedik) supported by COST (European Cooperation in Science and Technology).

---

**Table 1.**
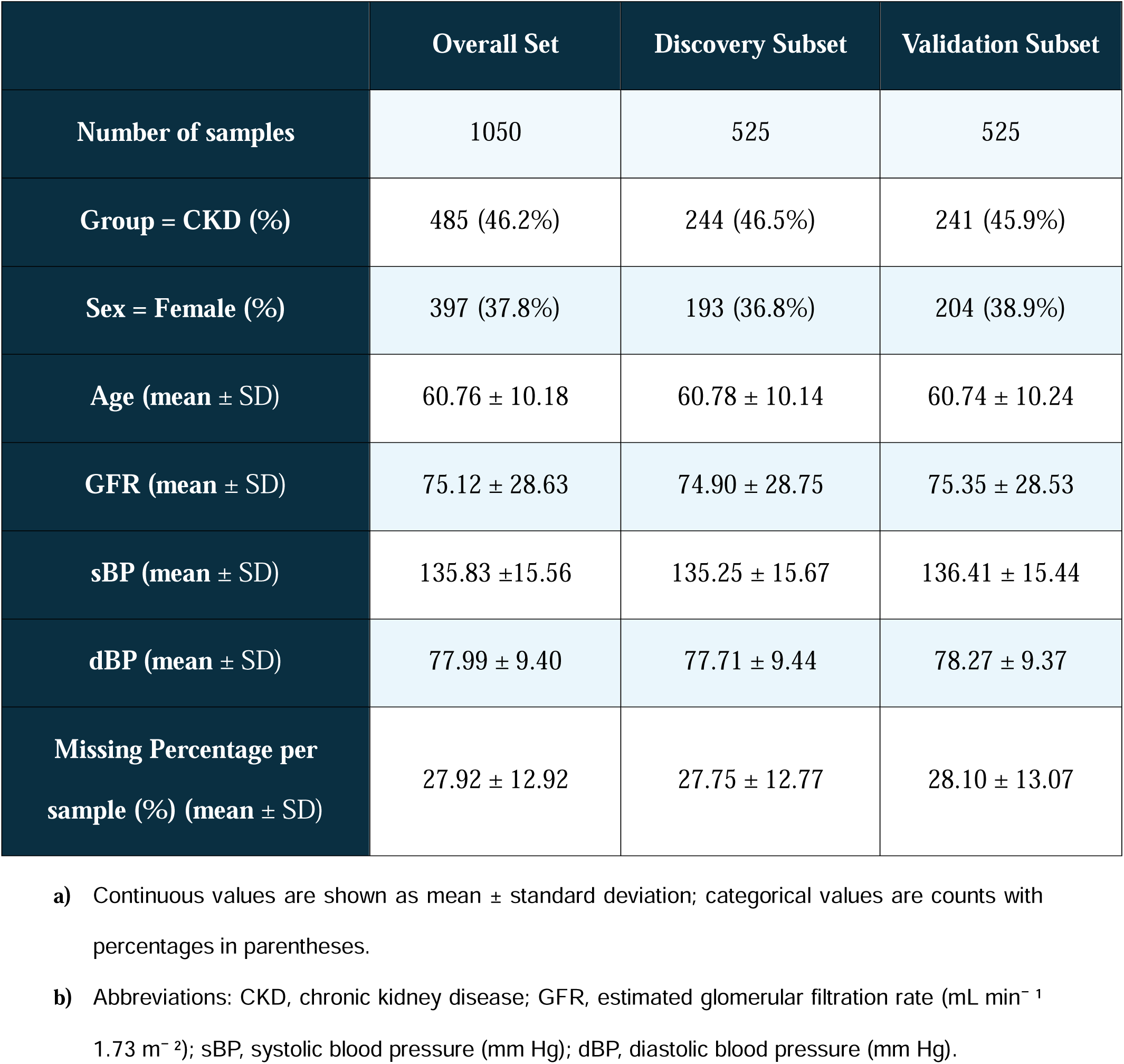
Clinical, demographic and data-quality characteristics of the urine-peptidomics cohort. The 1,050 samples were randomly split 1:1 into a discovery (training) set (n = 525) and a validation (test) set (n = 525), preserving the distribution of key clinical variables.

