## Supplementary figures and images for "Comparative evaluation of imputation and batch-effect correction for proteomics/peptidomics differential-expression analysis"

### Supplementary Figure S1

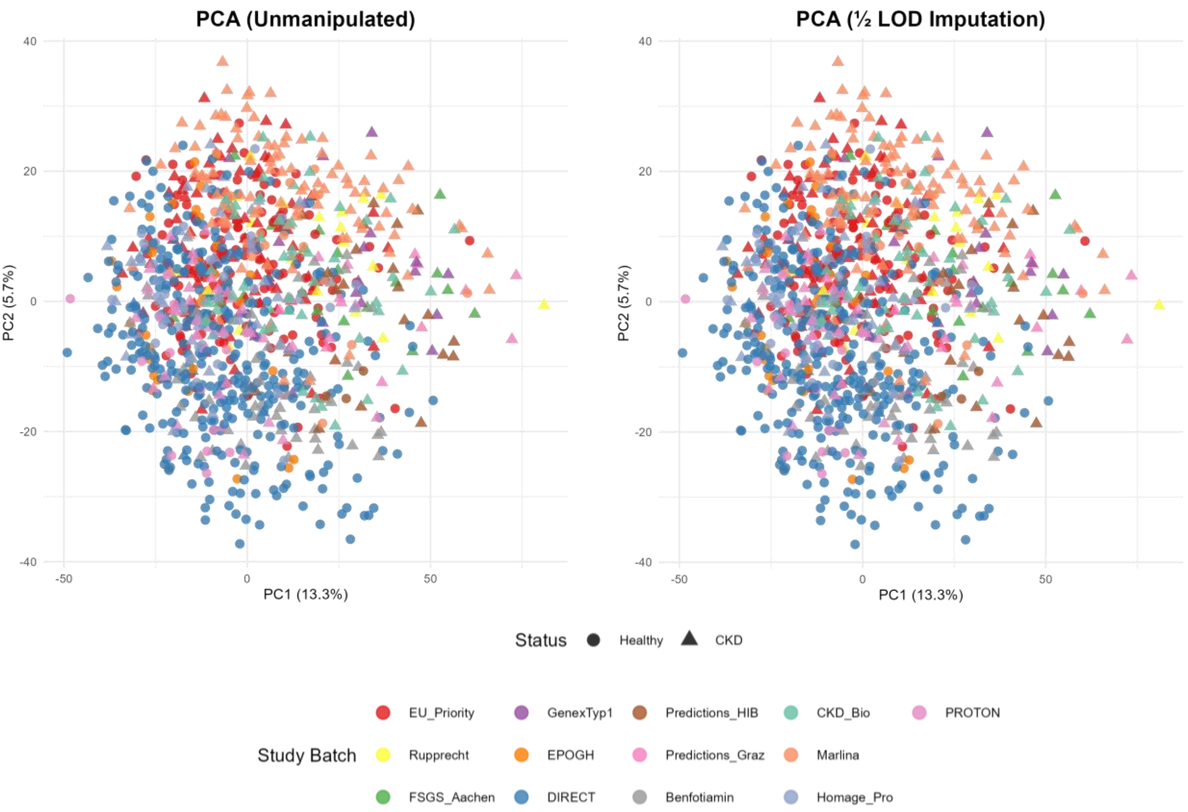

### Supplementary Figure S2

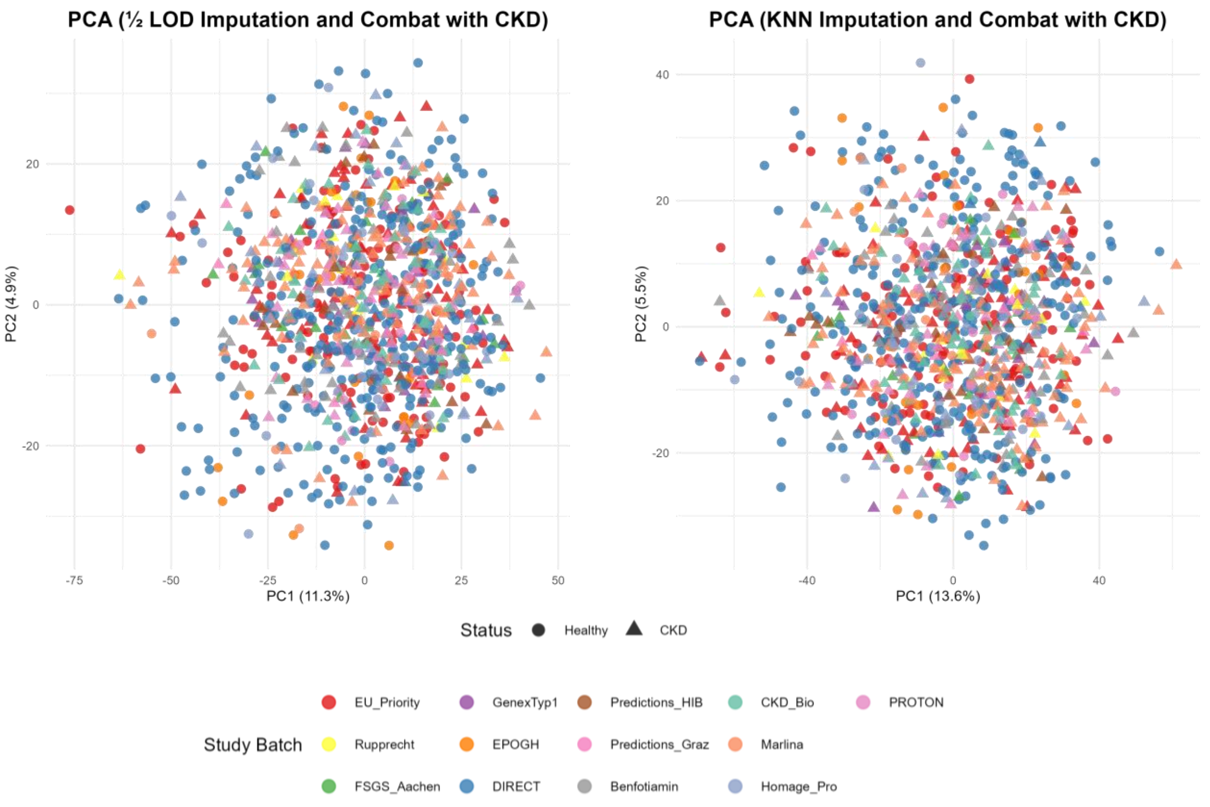
